# Prevalence and Characteristics of Steatotic Liver Disease in Germany - Magnetic Resonance Imaging in the German National Cohort (NAKO)

**DOI:** 10.64898/2026.05.29.26354407

**Authors:** Marc-Nicolas v. Itter, Elena Grune, Tobias Nonnenmacher, Stefan Rach, Martyna Flis, Tobias Haueise, Jakob Weiß, Hermann Brenner, Thomas Keil, Michael Roden, Matthias B. Schulze, Jeanette Schulz-Menger, Henry Völzke, Norbert Stefan, Christopher L. Schlett, Hans-Ulrich Kauczor, Jürgen Machann, Fabian Bamberg, Johanna Nattenmüller, Tobias Norajitra, Susanne Rospleszcz

## Abstract

**Background and Aims:** Steatotic liver disease (SLD) has high clinical and public health relevance. Robust population estimates of SLD and its subcategories are challenging due to the limitations of ultrasound measurements or non-invasive scores, particularly for low-grade steatosis. We aimed to quantify SLD prevalence using magnetic resonance imaging (MRI) in the population-based German National Cohort (NAKO).

**Methods:** Hepatic multi-echo Dixon MRI was performed at 5 dedicated study sites with identical setup across Germany. Liver fat (proton density fat fraction, PDFF), R2* as proxy for liver iron, and liver volume were assessed. The resulting data of N = 29’842 individuals (age range 20-72 years) were weighted by survey weights for regional representativeness, resulting in a sample of 50% women and a mean age of 45.6 years. SLD was defined as PDFF ≥ 5.75%, and sex-specific prevalence according to age, BMI, socioeconomic status and geographic region was calculated.

**Results:** Overall, SLD prevalence was 21.3% in women and 35.7% in men, and the majority were metabolic dysfunction-associated (MASLD, 89.3% of all SLD cases). Prevalence increased with age in a sex-specific pattern, suggesting potential menopausal effects in women. There was a relevant prevalence of SLD in individuals with normal weight (5.3% in women, 13.2% in men) and the age group <25 years (7.5% in women, 11.9% in women). Differences in prevalence between low and high socioeconomic status were more pronounced in women (37% vs 15.8%) compared to men (45.5% vs 30.3%).

**Conclusions:** Data underscore the high public health relevance of SLD and its subcategory MASLD. The considerable prevalence in groups historically considered low-risk, such as younger or lean individuals, emphasizes the need for raising awareness early.

## Introduction

Steatotic liver disease (SLD) represents a major public health burden, affecting approximately 25-33% of the global population [1]. Early stages are mainly asymptomatic, but SLD can advance to metabolic-associated steatohepatitis (MASH), liver cirrhosis, and hepatocellular carcinoma, and thus represent a substantial morbidity burden [2], as well as an increased burden of CVD.

The term non-alcoholic fatty liver disease (NAFLD) has traditionally been used to describe hepatic steatosis in the absence of relevant alcohol consumption. However, this terminology inadequately reflects the strong association between hepatic steatosis and metabolic dysfunction, including obesity, type 2 diabetes, insulin resistance, and dyslipidemia. To better capture this pathophysiological context and to avoid stigmatizing terminology, the concept of NAFLD has been renamed metabolic dysfunction–associated steatotic liver disease (MASLD), with an updated definition [3].

In parallel with this nomenclature update, other subcategories of SLD were reclassified. In addition to MASLD, metabolic dysfunction–associated alcohol-related liver disease (MetALD) was defined as MASLD with moderate alcohol consumption, whereas alcohol-related liver disease (ALD) was redefined based on higher alcohol intake thresholds [3, 4].

Robust estimations of population prevalences of SLD and its subcategories are difficult to establish, since dedicated scores, such as the fatty liver index (FLI) or the hepatic steatosis index (HSI) only have moderate diagnostic validity [5]. While invasive liver biopsy remains the gold standard to diagnose steatohepatitis and fibrosis, the current reference standard to assess SLD non-invasively is magnetic resonance imaging (MRI), which provides quantitative measurements even for non-pathologic fat content. In particular, MRI-based proton density fat fraction (PDFF) assessed by multi-echo Dixon sequences shows excellent agreement with magnetic resonance spectroscopy and histological fat quantification also in low-grade steatosis, while avoiding the sampling error inherent to liver biopsy [6]. In addition, measures of liver volume and proxies of liver iron – a known risk factor for SLD progression and advanced liver disease – can be simultaneously quantified with MRI [7].

A detailed understanding of the prevalence and population-level distribution of SLD is essential to support preventive strategies, including lifestyle-based [8] or medication-based [9] interventions. SLD is very prevalent in individuals living with overweight or obesity, however it can also occur in individuals with normal weight [10] and in young individuals [11], necessitating analyses in well-characterized cohorts with a wide range in age and body mass index (BMI).

The German National Cohort (NAKO), offers the opportunity to study population-based MRI data for individuals aged 20-72 years [12], weighted for regional representativeness [13]. Advances in automated image analysis and segmentation algorithms now enable the large-scale extraction of high-quality quantitative imaging biomarkers from these datasets to assess liver health. In the present study, we aim to quantify the prevalence of SLD and its subcategories – MASLD, MetALD, and ALD – based on MRI-derived hepatic fat fraction in this German population-based study. We characterize prevalences according to age, BMI, geographic region, and socioeconomic status. Furthermore, we investigate additional characteristics of liver health, such as liver volume, iron, and enzymes. By providing population-level data, this study establishes a robust resource for understanding the epidemiology of SLD in Germany and supports future prevention and research efforts.

## Methods

### Study sample

The population-based NAKO study recruited more than 205’000 participants in 18 study centers in Germany [14]. Of those, 30’868 participants from 11 study centers underwent whole-body MRI at five dedicated study sites. Details on inclusion criteria for the MRI examination and the imaging protocol have been reported previously [12]. The current analysis is based on all participants with valid hepatic fat measurements (N = 29’842).

### MRI examination

MRI examinations were performed on study-dedicated 3T MR scanners (MAGNETOM Skyra, Siemens Healthineers, Erlangen, Germany) with identical hardware and software configurations at the five study sites. All sites applied the same standardized MRI protocol under local supervision of board-certified radiologists. For liver assessment, a multi-echo-Dixon 3D volume interpolated breathhold examination (VIBE) sequence with six echo times (1.23, 2.46, 3.69, 4.92, 6.15, and 7.38 ms) was acquired in axial orientation with a TR of 4.36ms, a flip angle of 9°, section thickness of 4.0 mm and an in-plane voxel size of 1.6 × 1.6 mm [15].

### Assessment of liver fat content, hepatic iron, and liver volume

Automated liver segmentation was performed using a nnU-Net–based deep learning model, initially trained on MRI data from the HELENA trial and subsequently adapted to the NAKO study through manual review and correction of candidate segmentations. Using this pipeline, all multi-echo Dixon image datasets were segmented automatically, quantifying hepatic fat content as PDFF in %, estimating hepatic iron content from the R2* maps as relaxation rate in 1/s, and quantifying total liver volume as ml. Liver volume was additionally indexed by body surface area for further statistical analysis.

### Definition of SLD and subcategories

SLD was defined as PDFF ≥ 5.75%, following a recent meta-analysis [16]. Additionally, we present data on other currently discussed cutoffs for MRI-derived hepatic fat content: 5.00%, 5.56%, and 6.4% [17, 18].

The subcategories MASLD, MetALD, ALD and other were defined according to established criteria and current national guidelines [3, 4]. Cardiometabolic risk factors were defined as Obesity: BMI ≥25 kg/m^2^ or a waist circumference >94 cm in men or >80 cm in women, Diabetes: HbA1c ≥ 5.7%, or a diagnosis/treatment of type 2 diabetes, Hypertension: elevated blood pressure (≥130/85 mmHg) or antihypertensive treatment, High Cholesterol: plasma triglycerides ≥1.70 mmol/L or lipid-lowering therapy, and Low HDL-cholesterol (≤1.0 mmol/L in men, ≤1.3 mmol/L in women) or lipid-lowering therapy [3]. Daily alcohol consumption was calculated based on self-reported type and quantity of alcoholic beverages consumed. MASLD was defined as SLD with at least one cardiometabolic risk factor and low alcohol consumption (<20 or <30 g/day in women or men, respectively). MetALD was defined as SLD with at least one cardiometabolic risk factor and moderate alcohol consumption (<50 or <60 g/day in women or men, respectively). ALD was defined as SLD with high alcohol consumption (≥50 or ≥60 g/day in women or men, respectively). All individuals with SLD who met the criteria for neither MASLD, MetALD or ALD were subsumed in the subcategory “other”, which might indicate e.g. cryptogenic SLD, monogenic disease, or other etiologies such as drug-induced liver injury.

### Covariates

Participants underwent standardized touchscreen questionnaires, interviews and physical examinations at the respective NAKO study centers [14] (Supplementary Text S1). Clinical chemistry was done according to standard laboratory practice (Supplementary Text S1). Socioeconomic status was based on years of schooling and educational and vocational qualification according to the standardized ISCED categorization [19]. Geographic region was categorized into East (study centers Berlin-North, Berlin-South, Berlin-center, Neubrandenburg), North (study centers Düsseldorf, Essen, Münster), and South (study centers Augsburg, Freiburg, Mannheim, Saarbrücken).

### Statistical methods

Survey weights were constructed to approach representativeness of the sample with respect to the study regions, using population marginals of the official German microcensus at the level of administrative districts [13]. Calibration was based on age-group, sex, nationality (German vs. non-German), education, household size (1 vs 2 vs >=3), and migration background. Further details are described in [13].

Sample characteristics are presented as weighted mean and standard deviation, or weighted median with first and third quartile for continuous covariates, and weighted percentage for categorical covariates. The prevalence of SLD is given according to PDFF cutoffs stated above according to age, BMI, socioeconomic status and geographic region. Data are visualized by smoothed LOESS plots or bar charts. We used R version 4.5.1 with package “survey” for analyses.

## Results

### Study population

After weighting, the sex distribution in the sample was completely balanced (Figure S1). Mean age was 45.6 years, and 46% of women were post menopause (Table 1). Mean BMI was 26.6 kg/m^2^. 12.9% of women, but only 10.2% of men had low socioeconomic status, as defined by education years. Only 1.2% of women and 3.5% of men reported high alcohol consumption (Table 1). Regarding metabolic risk factors according to MASLD criteria, 81.6% (78.2% of women and 85.2% of men) had at least one metabolic risk factor, with obesity being the most prevalent (63% prevalence in women and 65.2% in men, Table 2).

**Table 1:** Sociodemographic characteristics and risk factor profile of the study population, weighted.

|  | Overall | Women | Men |
| --- | --- | --- | --- |
| % of total sample | 100 | 50 | 50 |
| **Sociodemographics** |  |  |  |
| Age, years | 45.6 (13.9) | 46.8 (14.1) | 44.4 (13.7) |
| Post menopause |  | 46 |  |
| Geographic region |  |  |  |
| East | 40.8 | 40.8 | 40.9 |
| North | 19.3 | 19.9 | 18.7 |
| South | 39.8 | 39.2 | 40.5 |
| Socioeconomic status |  |  |  |
| low | 11.6 | 12.9 | 10.2 |
| medium | 56.4 | 57.4 | 55.4 |
| high | 32.0 | 29.6 | 34.4 |
| **Anthropometry** |  |  |  |
| Body height, cm | 171.7 (9.6) | 165.0 (6.8) | 178.3 (7.2) |
| Body weight, kg | 78.6 (17.1) | 71.5 (15.4) | 85.8 (15.6) |
| BMI, kg/m<sup>2</sup> | 26.6 (5.2) | 26.3 (5.7) | 27.0 (4.6) |
| BMI category |  |  |  |
| Underweight | 1.3 | 1.7 | 0.9 |
| Normal weight | 41.8 | 48 | 35.7 |
| Overweight | 35.5 | 29.2 | 41.8 |
| Obesity class 1/2 | 18.9 | 17.9 | 20 |
| Obesity class 3 | 2.4 | 3.2 | 1.6 |
| **Medication intake** |  |  |  |
| Lipid-lowering | 7.2 | 6.5 | 7.9 |
| Glucose-lowering | 3.9 | 3 | 4.8 |
| Antihypertensive | 19.5 | 19.2 | 19.8 |
| **Behavioral risk factors** |  |  |  |
| Smoking behaviour |  |  |  |
| Never | 45.7 | 50.5 | 40.9 |
| Former | 29.5 | 27.1 | 32 |
| Current | 24.8 | 22.5 | 27.1 |
| Alcohol consumption |  |  |  |
| None | 11.0 | 11.6 | 10.4 |
| Low: <20 (women) or 30 (men) g/day | 79.8 | 81.7 | 78 |
| Moderate: <50 (women) or 60 (men) g/day | 6.8 | 5.5 | 8.1 |
| High: ≥50 (women) or 60 (men) g/day | 2.4 | 1.2 | 3.5 |
| **Cardiometabolic risk factors for SLD subcategory definition** |  |  |  |
| Obesity | 64.1 | 63 | 65.2 |
| Diabetes | 6.2 | 5.4 | 7 |
| Low HDL Cholesterol | 15.8 | 16.9 | 14.6 |
| High Cholesterol | 45.3 | 39.3 | 51.3 |
| Hypertension | 50.3 | 41.1 | 59.5 |
| Sum of risk factors (min zero to max 5) |  |  |  |
| 0 | 18.4 | 21.8 | 14.8 |
| 1 | 25.4 | 28.1 | 22.8 |
| 2 | 26.1 | 25 | 27.1 |
| 3 | 20.9 | 17.1 | 24.8 |
| 4 | 7.6 | 6.3 | 8.9 |
| 5 | 1.7 | 1.7 | 1.6 |

Data are weighted mean (standard deviation) for continuous variables, and weighted percentage for categorical variables. BMI categories: underweight: <18.5 kg/m<sup>2</sup>, normal weight: [18.5, 25.0) kg/m<sup>2</sup>, overweight: [25.0, 30.0) kg/m<sup>2</sup>, Obesity class 1/2: [30.0, 40.0) kg/m<sup>2</sup>, Obesity class 3: above 40 kg/m<sup>2</sup>. Note that there is no exact correspondence to the obesity definition for SLD subcategorization, as this one additionally includes waist circumference.

**Table 2:** Hepatic characteristics in the study population, weighted.

|  | Overall | Women | Men |
| --- | --- | --- | --- |
| SLD: PDFF ≥ 5.75% | 28.5 | 21.3 | 35.7 |
| MASLD | 25.0 | 19.7 | 30.4 |
| MetALD | 2.3 | 1.2 | 3.4 |
| ALD | 1.1 | 0.5 | 1.8 |
| PDFF, % (median [Q1, Q3]) | 3.4 [2.3, 6.6] | 2.8 [2.0, 4.9] | 4.0 [2.8, 8.2] |
| Liver volume, ml | 1613.3 (386.9) | 1453.3 (321.5) | 1773.5 (380.5) |
| Liver volume, ml/m<sup>2</sup> | 839.8 (135.8) | 813.0 (128.6) | 866.7 (137.4) |
| R2* as proxy for liver Iron, 1/s (median [Q1, Q3]) | 43.9 [37.9, 53.9] | 40.0 [35.4, 47.1] | 48.8 [41.7, 59.7] |
| **Liver enzymes** |  |  |  |
| ASAT, UL (median [Q1, Q3]) | 22.2 [18.0, 26.4] | 19.8 [16.8, 24.0] | 24.0 [19.8, 28.8] |
| ALAT, UL (median [Q1, Q3]) | 25.8 [19.8, 34.8] | 22.2 [18.0, 28.2] | 31.8 [24.6, 42.0] |
| GGT, UL (median [Q1, Q3]) | 25.8 [19.2, 37.2] | 21.6 [16.8, 28.8] | 31.2 [23.4, 45.6] |
| **Other** |  |  |  |
| Self-reported Hepatitis B | 1.3 | 1.2 | 1.4 |
| Self-reported Hepatitis C | 0.4 | 0.4 | 0.4 |

Data are weighted mean (standard deviation) or weighted median [Q1, Q3] for continuous variables, and weighted percentage for categorical variables. SLD: steatotic liver disease, PDFF: proton density fat fraction

### Prevalence of SLD, and characteristics of hepatic health

Median hepatic PDFF was 2.8% [2.0%, 4.9%] in women and 4.0% [2.8%, 8,2%] in men (Table 2). Overall prevalence of SLD, as defined by a PDFF ≥ 5.75%, was 21.3% in women and 35.7% in men, with the majority being MASLD (25.0% population prevalence, corresponding to 89.3% of all SLD cases). A more lenient cutoff of 5.00% resulted in a SLD prevalence of 24.3% in women and 39.8% in men, whereas a stricter cutoff of 6.4% resulted in a SLD prevalence of 18.8% in women and 32.4% in men (Supplementary Table S1). Liver volume and iron, as well as liver enzymes were lower in women compared to men (Table 2).

### Distribution of SLD according to age, BMI, geographic region and socioeconomic status

In both women and men, prevalence of SLD increased with age, but age-related patterns differed substantially (Figure 1A). In men, prevalence increased almost linear with age until saturation around age 60, while prevalence in women increased steeply only after age 50 (Figure 1A). Importantly, there was a relevant prevalence already in the younger age groups: Prevalence in the age group 20-24 years was 7.5% in women and 11.9% in men (Supplementary Tables S3 and S4).

**Figure 1:**
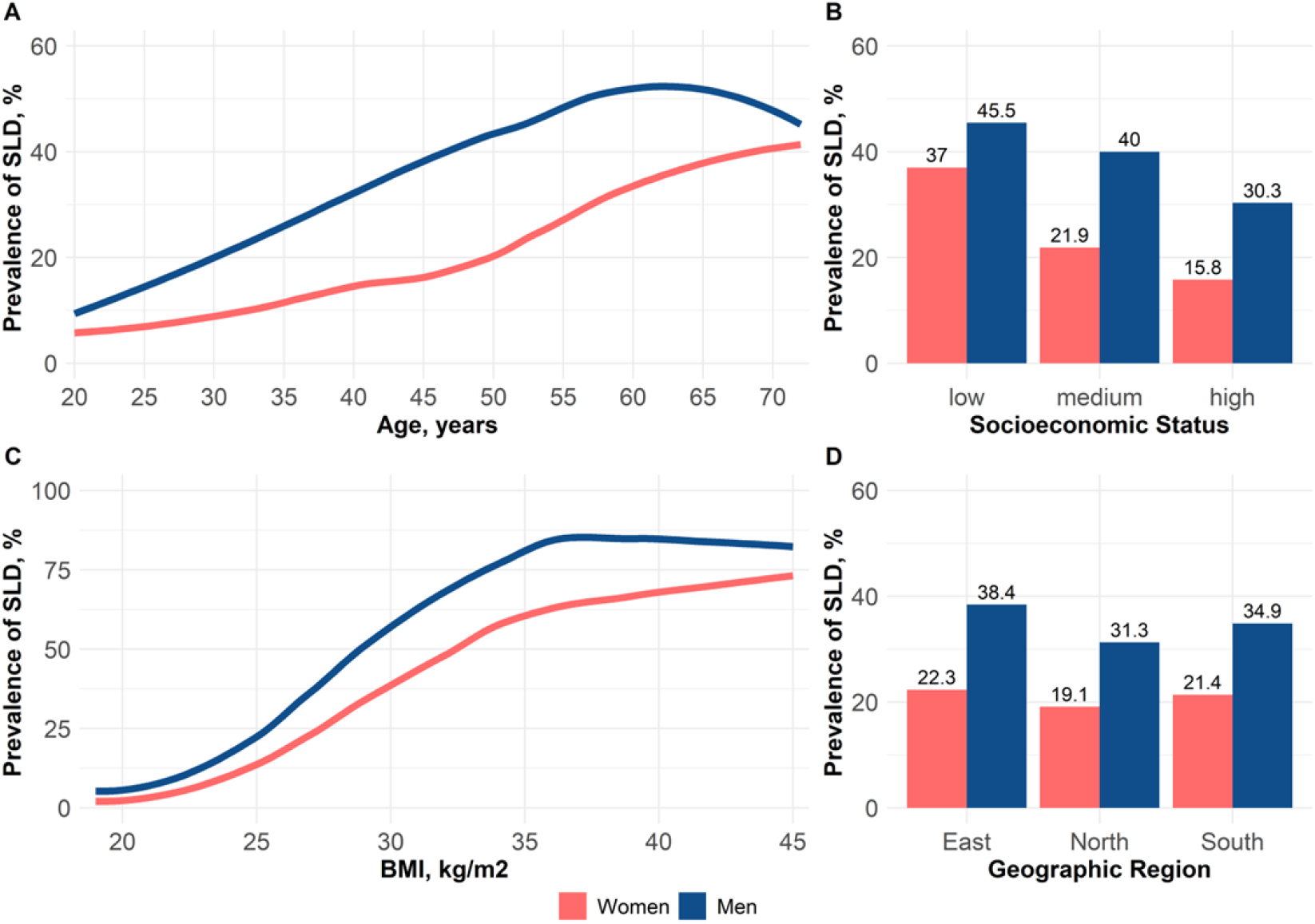
Prevalence of SLD, defined by PDFF≥5.75% On the x-axis: continuous age (A) or BMI (C), or categories of socioeconomic status (B) and geographic region (D). On the y-axis: Prevalence of SLD for women (red) and men (blue). Graphs according to continuous age and BMI are smoothed with LOESS.

The prevalence of SLD was higher in individuals with low socioeconomic status compared to those with medium or high socioeconomic status (Figure 1B). However, this gradient was stronger in women than in men. In women with low socioeconomic status, SLD prevalence was more than double compared to women with high socioeconomic status (37% vs 15.8%), whereas in men with low socioeconomic status, SLD prevalence was only 1.5-fold compared to men with high socioeconomic status (45.5% vs 30.3%, Figure 1B).

Prevalence patterns according to BMI were similar in women and men (Figure 1C), with steep increases in SLD prevalence with higher BMI. Importantly, there was a relevant prevalence already in individuals with BMI corresponding to normal weight: The prevalence in this group (BMI between 18.5 and 25 kg/m^2^) was 5.3% in women and 13.2% in men (Supplementary Tables S5 and S6). Regarding geographic region, prevalence was highest in East Germany, followed by South, and lowest in North in both, women and men (Figure 1D). All patterns and trends were similar for different cutoffs for SLD definition (Supplementary Figures S2-S4).

### Distribution of SLD subcategories

MASLD accounted for the majority of SLD in both women (19.7% population prevalence, corresponding to 92.5% of all SLD cases, Table 2) and men (30.4% population prevalence, corresponding to 85.2% of all SLD cases, Table 2). Consequently, MASLD prevalence patterns according to age, socioeconomic status, BMI and geographic region mirrored those of SLD (Figure 2). The prevalence of MetALD and ALD was substantially higher in men than in women (Table 2). These subcategories were most prevalent in men with low socioeconomic status, and men in the geographic region South Germany (Figure 2). Prevalence of the subcategory “other” was negligible in both women and men (prevalence <0.01%).

**Figure 2:**
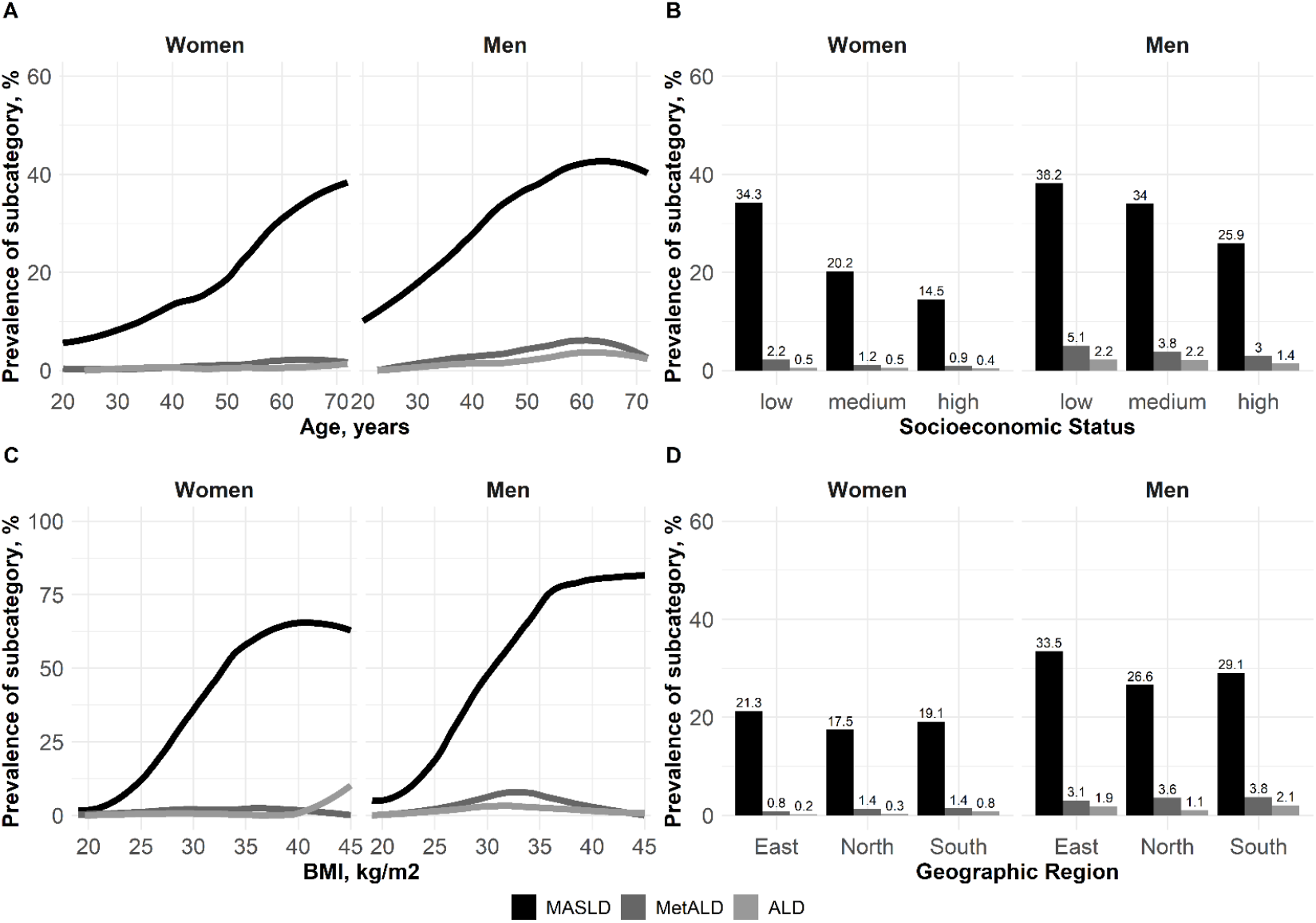
Prevalence of SLD subcategories On the x-axis: continuous age (A) or BMI (C), and categories of socioeconomic status (B) or geographic region (D). On the y-axis: Prevalence of SLD subcategory for women (left facet) and men (right facet). Graphs according to continuous age and BMI are smoothed with LOESS. Subcategory “other” not shown due to negligible prevalence.

### Characteristics of liver health in SLD

Individuals with SLD were more likely to have at least one metabolic risk factor (97.9% vs 75.2% in individuals without SLD), with obesity being the most prevalent (92.3% vs 52.9%, Supplementary Table S2). Individuals with SLD had higher median hepatic PDFF (11.0% [7.5%, 16.6%] vs 2.7% [2.1%, 3.6%], Supplementary Table S2), and higher liver volume, iron, and enzymes (e.g. GGT: 36.6 UL vs 23.4 UL, Table S2).

Furthermore, age-related patterns of hepatic PDFF, iron, volume and enzymes differed between individuals with and without SLD (Figure 3). Hepatic PDFF increased more steeply with age in individuals with SLD (Figure 3A). Hepatic iron showed similar age patterns in women with and without SLD, with steep increases during the age range consistent with menopausal transition (Figure 3B). However, iron levels increased more steeply with age in men with SLD compared to men without SLD. This might be explained by the higher prevalence of MetALD and ALD in men, since R2* as a proxy for liver iron was higher in these alcohol-related subcategories (Supplementary Figure S5). Notably, the liver enzyme ALAT was substantially increased in younger men with SLD (Figure 3E).

**Figure 3:**
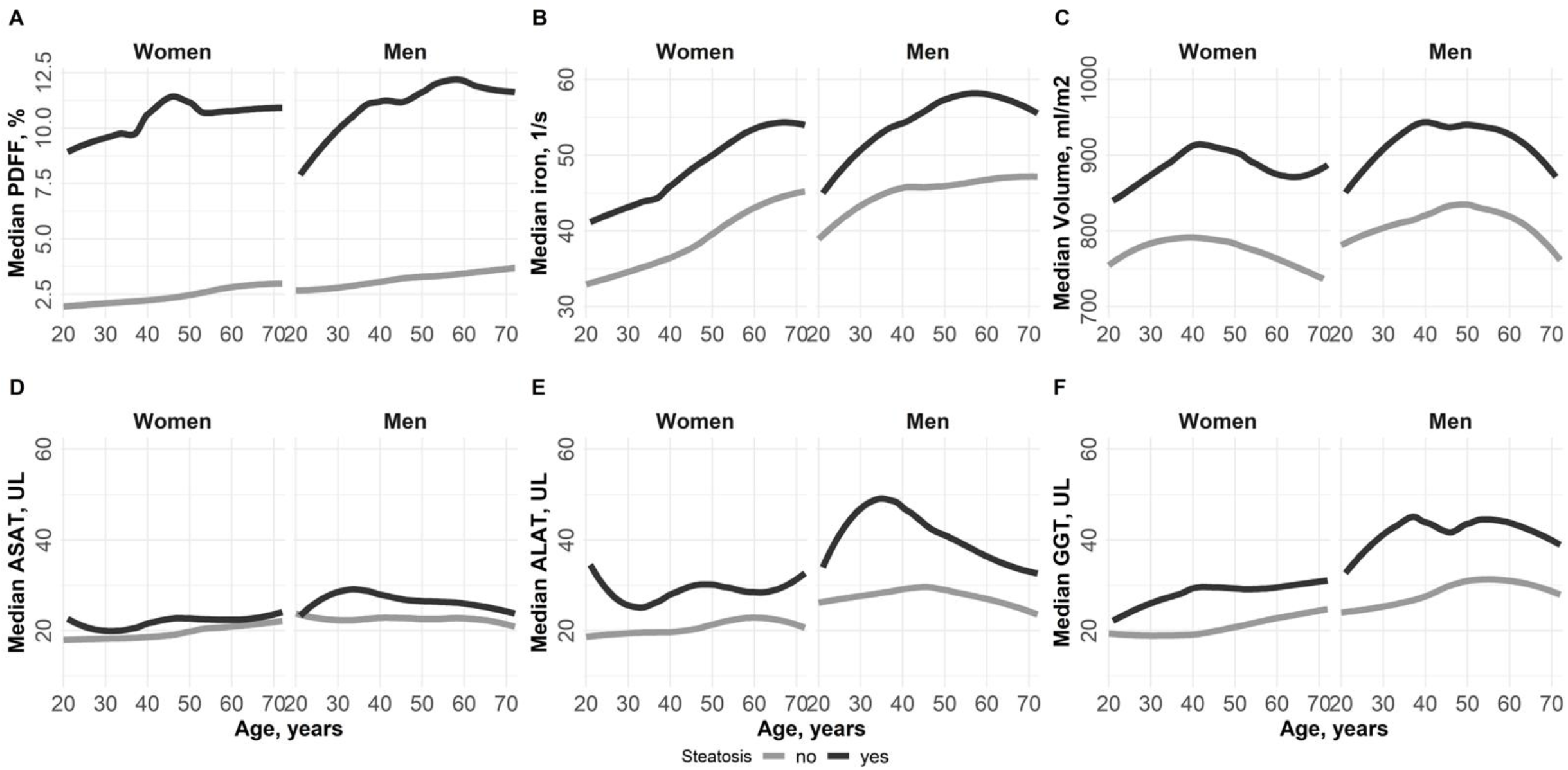
Hepatic characteristics according to presence of SLD On the x-axis: continuous age. On the y-axis: Median value of liver fat (A), R2* as a proxy for liver iron (B), liver volume, indexed by body surface area (C), enzyme ASAT (D), enzyme ALAT (E) and enzyme GGT (F) for women (left facet) and men (right facet) according to presence of SLD. Graphs are smoothed with LOESS.

## Discussion

In this large, weighted, population-based study, we report the prevalence of MRI-derived SLD and its subcategories stratified by sex, age, geographic region, BMI and socioeconomic status, and accompanying values of liver volume, hepatic iron content and liver enzymes.

We observed a high prevalence of SLD, with an overall prevalence up to 39.8% in men, and up to 24.3% in women, depending on the liver fat cutoff used. The majority of cases was MASLD, which is in line with other large ultrasound studies from the US [20] or Korea [21]. However we cannot exclude that the low prevalence of MetALD and ALD might also partly be due to underreporting of alcohol consumption.

Our findings indicate that the prevalence of SLD and MASLD increases with age, with an initial steeper slope in men and a more significant increase in prevalence among women in the postmenopausal age range. This discrepancy in the development of SLD between men and women can likely be partially attributed to the influence of reproductive hormones. In studies still using previous nomenclature, not only the prevalence, but also the disease severity was shown to increase after menopause [22], and NAFLD and NASH were the primary indications for liver transplantation in women without hepatocellular carcinoma [23]. In our cohort, the higher MASLD prevalence observed in women in the postmenopausal age range also reflect menopause-associated increases in visceral adiposity and dysregulation of carbohydrate and lipid metabolism [22].

Our data show that the prevalence of SLD increased steadily with age, underscoring aging as a major risk factor not only for disease onset but also for disease progression and complications [24]. Also, aging is frequently accompanied by sarcopenia, which has been independently linked to SLD: Individuals with progressive muscle loss have a 1.5-to 2.5-fold higher risk of hepatic steatosis, highlighting the interplay between muscle mass, metabolism, and liver health [25]. However, the considerable prevalence of SLD in the younger age groups (7.5% in women and 11.9% in men aged 20-24 years) emphasizes the need for raising awareness and prevention efforts early on.

As expected, SLD prevalence increased with higher BMI, confirming obesity as a major risk factor for hepatic steatosis [26]. However, there was a considerable prevalence of SLD in individuals with a BMI indicating normal weight (5.3% in women and 13.2% in men). Moreover, a subgroup of individuals with elevated BMI did not exhibit hepatic steatosis, highlighting the limitations of BMI as a surrogate for metabolic risk. Generally, different body composition patterns relate to different degrees of cardiometabolic risk [27]. BMI does not capture fat distribution, and visceral adipose tissue confers a substantially higher risk for SLD than subcutaneous fat [28], which underscores the need for more refined measures of adiposity beyond BMI alone.

We further observed marked geographic differences in SLD prevalence, with the highest prevalence in Eastern Germany, followed by Southern Germany, and the lowest prevalence in northern regions. The increased burden in Eastern Germany may be partly explained by regional disparities in healthcare infrastructure [29] and a higher prevalence of cardiometabolic risk factors, especially obesity and diabetes, as previously reported [30]. Although our results have been derived from German data, geographic disparities in liver health outcomes transfer to other countries as well [31], highlighting the complex interplay between regional deprivation, socioeconomic status and health behavior.

In line with this, the prevalence of SLD was particularly high in individuals with a low socioeconomic status (SES), assessed using education-based ISCED-97 categories, supporting earlier findings [32]. The difference of SLD prevalence between low and high SES was more pronounced in women than in men, and more pronounced for MASLD, suggesting that social determinants of health play a critical role in metabolically driven liver disease. The greater vulnerability of women with low SES is consistent with evidence linking low SES to higher cardiovascular risk among women compared with men [33]. Conceptually, a lower SES can influence unhealthier diet choices [34] and thus favor the development of SLD and especially its subcategory MASLD. A study from the UK showed that lower food expenditure in lower SES groups resulted in less-healthy food choices [35].

In our cohort, R2* as a proxy for liver iron was higher in individuals with SLD, and elevated in women in the post-menopausal age range. Hepatic iron overload is a recognized risk factor for SLD progression and advanced liver disease [7]. The coexistence of elevated liver iron and SLD may accelerate disease progression through multiple mechanisms, including insulin resistance-associated hyperinsulinemia, increased ferritin levels, and reduced hepatic hepcidin synthesis, resulting in systemic iron accumulation [36]. In postmenopausal women, the cessation of menstrual blood loss represents an additional contributor to increased iron stores [37]. We furthermore observed a gradual increase of R2* as a proxy for liver iron in MetALD and ALD, reflecting a further adverse interaction between alcohol consumption and iron metabolism. These findings support previous research from population-based imaging that showed associations between alcohol consumption and increased liver iron for both men and women [38]. Since both alcohol and iron induce oxidative stress, lipid peroxidation, and hepatocellular injury, their interplay can accelerate fibrosis progression and increase the risk of cirrhosis [39].

Biochemical liver markers supported the imaging findings. Individuals with SLD exhibited higher liver enzyme levels, with ALAT showing a more pronounced increase than ASAT, particularly in men. This pattern is consistent with observations from the development of the Framingham Steatosis Index, where the ALAT/ASAT ratio provided superior discrimination for hepatic steatosis compared with either enzyme alone [40]. Importantly, normal ALAT values do not exclude SLD, highlighting the limited sensitivity of single enzyme thresholds [41].

In conclusion, our data provide population-level estimates of SLD and its subcategories in Germany, with detailed stratification by demographic, anthropometric, geographic, and socioeconomic factors. The use of six-point multi-echo Dixon MRI – considered equivalent to magnetic resonance spectroscopy as a non-invasive reference standard – enabled accurate whole-liver fat quantification while avoiding sampling bias inherent to ultrasound and histopathology. The application of statistical weighting, aiming for representativeness within the study regions, further enhances the generalizability of our findings. Nevertheless, it must be considered that the cross-sectional design precludes assessment of longitudinal disease trajectories and individual progression of SLD. Furthermore, we could not account for potential underreporting of alcohol consumption, which may have caused an underestimation of MetALD and ALD prevalence.

Taken together, our findings establish an important epidemiological resource for further hepato-metabolic disease research, and underscore the relevance of SLD as a major public health concern.

## Data Availability

Access to and use of NAKO (German National Cohort) data and biosamples can be obtained via an electronic application portal (https://transfer.nako.de/transfer/index).

## Declarations

### CRediT author statement

Conceptualization: SR; Investigation: MNvI, EG, TNon, StR, MF, TH, TN, SR; Methodology: EG, SR; Formal analysis: EG, SR; Resources: MNvI, EG, TNon, StR, MF, TH, JW, HB, TK, MR, MBS, JSM, HV, NS, CLS, HUK, JM, FB, JN, TN, SR; Data Curation: EG, TNon, StR, TN, SR ; Visualization: EG, SR; Writing - original draft: MNvI, SR; Writing - review and editing: MNvI, EG, TNon, StR, MF, TH, JW, HB, TK, MR, MBS, JSM, HV, NS, CLS, HUK, JM, FB, JN, TN, SR

## Acknowledgments

This project was conducted with data (application no. NAKO-751) from the German National Cohort (NAKO). We thank all participants who took part in the NAKO study and the staff of this research initiative.

