## Supplementary Material for "Prevalence and Characteristics of Steatotic Liver Disease in Germany - Magnetic Resonance Imaging in the German National Cohort (NAKO)"

v.Itter et al.

### Supplementary Texts

**Supplementary Text S1:** Covariate assessment

Information on previous Hepatitis B or C infection, smoking status, alcohol consumption, and use of antihypertensive, antidiabetic, and lipid-lowering medications was self-reported.

Body height and weight were measured in light clothing with a stadiometer 274 and a medical Body Composition Analyzer (mBCA) 515 (seca GmbH & Co. KG, Hamburg), respectively. BMI was calculated as weight in kg divided by squared height in meter. Body surface area (for indexation of liver volume) was calculated by duBois formula as 0.007184 x height in $cm^{0.725}$ x weight in $kg^{0.425}$. Waist circumference was measured with an inelastic tape (seca 201) halfway between the lowest rib and the iliac crest.

Blood pressure was measured on the right arm in seating position. After an initial waiting period of 5 minutes, 2 consecutive measurements were taken with 2 minutes in between. The second value was used as final.

Glucose, total cholesterol, LDL cholesterol, HDL cholesterol, and triglycerides were measured from serum by enzymatic photometry (Dimension VISTA 1500, Siemens Healthineers, Erlangen, Deutschland). HbA1c was measured from EDTA whole blood by HPLC (Tosoh G8 HPLC Analyzer, Tosoh Bioscience, Inc., San Francisco, USA).

Menopause was defined as age greater than 60 years, no menstrual bleeding for more than 6 consecutive months in women aged 45 years or older or permanent cessation of menstruation due to illness.

ASAT, ALAT, GGT, and AP were measured in serum by enzymatic photometry (Dimension VISTA 1500, Siemens Healthineers, Erlangen, Deutschland).

### Supplementary Tables

**Supplementary Table S1:** Prevalence of steatotic liver disease and its subcategories by different cutoffs

| | Overall | Women | Men |
|---|---|---|---|
| SLD: PDFF ≥ 5.75% | 28.5 | 21.3 | 35.7 |
| MASLD | 25.0 | 19.7 | 30.4 |
| MetALD | 2.3 | 1.2 | 3.4 |
| ALD | 1.1 | 0.5 | 1.8 |
| SLD: PDFF ≥ 5% | 32.1 | 24.3 | 39.8 |
| MASLD | 28.1 | 22.4 | 33.8 |
| MetALD | 2.7 | 1.4 | 3.9 |
| ALD | 1.3 | 0.5 | 2.1 |
| SLD: PDFF ≥ 5.56% | 29.3 | 22 | 36.6 |
| MASLD | 25.7 | 20.3 | 31.2 |
| MetALD | 2.4 | 1.2 | 3.5 |
| ALD | 1.2 | 0.5 | 1.9 |
| SLD: PDFF ≥ 6.4% | 25.6 | 18.8 | 32.4 |

| | | | |
|---|---|---|---|
| MASLD | 22.4 | 17.3 | 27.5 |
| MetALD | 2.1 | 1.1 | 3.2 |
| ALD | 1.1 | 0.4 | 1.7 |

**Supplementary Table S2:** Characteristics of the study sample, according to steatotic liver disease (PDFF ≥ 5.75%)

| | without SLD | With SLD |
|---|---|---|
| % of total sample | 71.5 | 28.5 |
| **Sociodemographics** | | |
| Age, years | 43.3 (13.8) | 51.3 (12.5) |
| Male sex | 45.0 | 62.5 |
| Geographic region | | |
| East | 39.8 | 43.6 |
| North | 20.2 | 16.9 |
| South | 40.0 | 39.5 |
| Socioeconomic status | | |
| low | 9.8 | 16.0 |
| medium | 55.5 | 58.6 |
| high | 34.7 | 25.5 |
| **Anthropometry** | | |
| Body height, cm | 171.6 (9.5) | 172.0 (10.0) |
| Body weight, kg | 73.9 (14.4) | 90.5 (17.4) |
| BMI, kg/m$^2$ | 25.0 (4.2) | 30.6 (5.4) |
| BMI category | | |
| Underweight | 1.6 | 0.5 |
| Normal weight | 54.2 | 10.9 |
| Overweight | 33.5 | 40.6 |
| Obesity class 1/2 | 9.8 | 41.8 |
| Obesity class 3 | 0.9 | 6.2 |
| **Medication intake** | | |
| Lipid-lowering | 5.2 | 12.3 |
| Glucose-lowering | 1.8 | 91 |
| Antihypertensive | 12.9 | 36.0 |
| **Behavioral risk factors** | | |
| Smoking behaviour | | |
| Never | 47.8 | 40.1 |
| Former | 27.2 | 35.4 |
| Current | 24.9 | 24.5 |
| Alcohol consumption | | |
| No consumption | 10.4 | 12.7 |
| Low: <20 (women) or 30 (men) g/day | 82.0 | 74.3 |
| Moderate: <50 (women) or 60 (men) g/day | 6.0 | 8.7 |
| High: ≥50 (women) or 60 (men) g/day | 1.6 | 4.2 |

| **Cardiometabolic risk factors for SLD subcategory definition** | | |
|---|---|---|
| Obesity | 52.9 | 92.3 |
| Diabetes | 3.0 | 14.3 |
| Low HDL Cholesterol | 10.9 | 27.9 |
| High Cholesterol | 35.4 | 69.9 |
| Hypertension | 40.5 | 74.9 |
| Sum of risk factors (zero to max 5) | | |
| 0 | 24.8 | 2.1 |
| 1 | 31.5 | 10.1 |
| 2 | 25.8 | 26.7 |
| 3 | 14.4 | 37.4 |
| 4 | 3.3 | 18.3 |
| 5 | 0.2 | 5.3 |
| **Hepatic Characteristics** | | |
| PDFF, % (median [Q1, Q3]) | 2.7 [2.1, 3.6] | 11.0 [7.5, 16.6] |
| Liver volume, ml | 1492.2 (276.6) | 1917.3 (451.8) |
| Liver volume, ml/m$^2$ | 800.2 (97.1) | 939.2 (165.0) |
| R2* as a proxy for liver iron, 1/s (median [Q1, Q3]) | 40.6 [36.3, 47.2] | 54.4 [47.2, 64.1] |
| ASAT, UL (median [Q1, Q3]) | 21.0 [17.4, 25.2] | 25.2 [20.4, 31.2] |
| ALAT, UL (median [Q1, Q3]) | 23.4 [19.2, 30.0] | 34.8 [27.0, 49.2] |
| GGT, UL (median [Q1, Q3]) | 23.4 [18.0, 31.2] | 36.6 [27.0, 55.8] |
| Self-reported Hepatitis B | 1.1 | 1.7 |
| Self-reported Hepatitis C | 0.3 | 0.6 |

Data are weighted mean (standard deviation) or weighted median [Q1, Q3] for continuous variables, and weighted percentage for categorical variables. BMI categories: underweight: <18.5 kg/m$^2$, normal weight: [18.5, 25.0) kg/m$^2$, overweight: [25.0, 30.0) kg/m$^2$, Obesity class 1/2: [30.0, 40.0) kg/m$^2$, Obesity class 3: above 40 kg/m$^2$

**Supplementary Table S3:** Women: Prevalence of steatotic liver disease and its subcategories by age groups.

| | Overall | [20,25) | [25,30) | [30,35) | [35,40) | [40,45) | [45,50) | [50,55) | [55,60) | [60,65) | [65,70) | >70 |
|---|---|---|---|---|---|---|---|---|---|---|---|---|
| % of total sample | 100 | 6 | 9.7 | 9.7 | 8.9 | 7.2 | 10.7 | 13.1 | 11.7 | 11.3 | 9.6 | 2 |
| SLD: PDFF ≥ 5.75% | 21.3 | 7.5 | 8 | 8 | 14.4 | 14.7 | 17.8 | 21.2 | 32.6 | 36.7 | 35.9 | 43.7 |
| MASLD | 19.7 | 7 | 8 | 7 | 13 | 13.5 | 16.7 | 19.1 | 30.9 | 33.9 | 32.3 | 41.8 |
| MetALD | 1.2 | 0.4 | 0 | 0.5 | 0.3 | 1.2 | 0.7 | 1.7 | 1 | 2.6 | 2.6 | 1.3 |
| ALD | 0.5 | 0 | 0 | 0.5 | 1.1 | 0 | 0.4 | 0.5 | 0.7 | 0.2 | 0.9 | 0.6 |
| SLD: PDFF ≥ 5% | 24.3 | 8 | 8.8 | 9.5 | 17.6 | 17.2 | 20.6 | 24.7 | 36.7 | 40.5 | 42 | 46.5 |
| MASLD | 22.4 | 7.6 | 8.7 | 8.5 | 16 | 15.9 | 19.2 | 22.3 | 34.5 | 36.9 | 37.7 | 44.5 |
| MetALD | 1.4 | 0.5 | 0.1 | 0.5 | 0.5 | 1.2 | 1 | 1.8 | 1.5 | 3.4 | 3.1 | 1.5 |
| ALD | 0.5 | 0 | 0 | 0.5 | 1.1 | 0 | 0.4 | 0.5 | 0.7 | 0.2 | 1.1 | 0.6 |
| SLD: PDFF ≥ 5.56% | 22 | 7.5 | 8.1 | 8.1 | 14.8 | 15.3 | 18.4 | 22.6 | 33 | 37.7 | 37.2 | 44.5 |
| MASLD | 20.3 | 7 | 8.1 | 7.1 | 13.5 | 14.1 | 17.1 | 20.5 | 31.3 | 34.8 | 33.1 | 42.7 |
| MetALD | 1.2 | 0.4 | 0 | 0.5 | 0.3 | 1.2 | 0.8 | 1.7 | 1 | 2.6 | 2.9 | 1.3 |
| ALD | 0.5 | 0 | 0 | 0.5 | 1.1 | 0 | 0.4 | 0.5 | 0.7 | 0.2 | 1.1 | 0.6 |
| SLD: PDFF ≥ 6.4% | 18.8 | 6.7 | 7.2 | 5.7 | 12.9 | 11.7 | 15.7 | 18.5 | 29.2 | 32.8 | 31.8 | 43.2 |
| MASLD | 17.3 | 6.3 | 7.1 | 5 | 11.5 | 10.5 | 14.7 | 16.5 | 27.6 | 30.2 | 28.4 | 41.6 |
| MetALD | 1.1 | 0.4 | 0 | 0.2 | 0.3 | 1.2 | 0.6 | 1.5 | 1 | 2.4 | 2.4 | 1 |
| ALD | 0.4 | 0 | 0 | 0.5 | 1.1 | 0 | 0.4 | 0.4 | 0.6 | 0.1 | 0.9 | 0.6 |

Subcategory “Other” not shown because of negligible prevalence.

**Supplementary Table S4:** Men: Prevalence of steatotic liver disease and its subcategories by age groups.

| | Overall | [20,25) | [25,30) | [30,35) | [35,40) | [40,45) | [45,50) | [50,55) | [55,60) | [60,65) | [65,70) | >70 |
|---|---|---|---|---|---|---|---|---|---|---|---|---|
| % of total sample | 100 | 6.5 | 10.6 | 13.3 | 9.8 | 9 | 11.3 | 12.6 | 9.8 | 8.5 | 7.2 | 1.3 |
| SLD: PDFF ≥ 5.75% | 39.8 | 11.9 | 21.5 | 25.1 | 31 | 40.7 | 46.5 | 48.6 | 54.6 | 59.8 | 55.4 | 68.1 |
| MASLD | 33.8 | 11.5 | 19.7 | 21.6 | 27.7 | 33.7 | 41.5 | 40.5 | 44.7 | 48.5 | 45.2 | 61.1 |
| MetALD | 3.9 | 0.4 | 0.8 | 2.7 | 2.2 | 4 | 3.8 | 5.3 | 6.3 | 6.7 | 6.7 | 4.8 |
| ALD | 2.1 | 0 | 1 | 0.8 | 1.2 | 2.9 | 1.2 | 2.8 | 3.6 | 4.5 | 3.5 | 2.2 |
| SLD: PDFF ≥ 5% | 36.6 | 11.1 | 19 | 22.9 | 28.2 | 36.9 | 42.3 | 44.6 | 51.6 | 56 | 50.2 | 65.8 |
| MASLD | 31.2 | 10.9 | 17.4 | 20 | 24.9 | 31.4 | 37.7 | 37.3 | 42.1 | 45.5 | 40.7 | 59.6 |
| MetALD | 3.5 | 0.2 | 0.6 | 2.5 | 2.1 | 3.6 | 3.5 | 4.6 | 6 | 6.3 | 6.1 | 4 |
| ALD | 1.9 | 0 | 1 | 0.4 | 1.2 | 2 | 1.1 | 2.6 | 3.5 | 4.2 | 3.3 | 2.2 |
| SLD: PDFF ≥ 5.56% | 35.7 | 10.7 | 18.3 | 21.6 | 27.3 | 36.4 | 40.8 | 43.7 | 50.9 | 55 | 49.6 | 63.7 |
| MASLD | 30.4 | 10.7 | 17.2 | 18.8 | 24.2 | 30.9 | 36.3 | 36.5 | 41.5 | 44.7 | 40.3 | 57.8 |
| MetALD | 3.4 | 0 | 0.6 | 2.5 | 1.9 | 3.4 | 3.4 | 4.6 | 6 | 6.2 | 6 | 3.7 |
| ALD | 1.8 | 0 | 0.5 | 0.4 | 1.2 | 2 | 1.1 | 2.6 | 3.4 | 4.1 | 3.2 | 2.2 |
| SLD: PDFF ≥ 6.4% | 32.4 | 8.2 | 16.7 | 19 | 25.2 | 32.1 | 37.8 | 39.6 | 47.1 | 51.5 | 45.7 | 52.1 |
| MASLD | 27.5 | 8.2 | 15.6 | 16.3 | 22.1 | 27 | 33.7 | 32.8 | 38.4 | 41.8 | 37.7 | 46.6 |
| MetALD | 3.2 | 0 | 0.6 | 2.4 | 1.9 | 3.2 | 3.1 | 4.4 | 5.4 | 5.7 | 5 | 3.5 |
| ALD | 1.7 | 0 | 0.5 | 0.3 | 1.2 | 1.9 | 0.9 | 2.3 | 3.3 | 4 | 2.9 | 2.1 |

Subcategory "Other" not shown because of negligible prevalence.

**Supplementary Table S5:** Women: Prevalence of steatotic liver disease and its subcategories by BMI category.

| | underweight | normal weight | overweight | Obesity class 1/2 | Obesity class 3 |
|---|---|---|---|---|---|
| % of total sample | 1.7 | 48 | 29.2 | 17.9 | 3.2 |
| SLD: PDFF ≥ 5.75% | 8.6 | 5.3 | 23.4 | 53.5 | 69.5 |
| MASLD | 6.8 | 4.5 | 21.3 | 50.9 | 65.5 |
| MetALD | 1.8 | 0.6 | 1.5 | 2.3 | 0.7 |
| ALD | 0 | 0.2 | 0.6 | 0.3 | 3.4 |
| SLD: PDFF ≥ 5% | 10 | 6.5 | 27.3 | 59.8 | 74.4 |
| MASLD | 8.2 | 5.4 | 24.8 | 56.8 | 70.3 |
| MetALD | 1.8 | 0.8 | 1.9 | 2.6 | 0.7 |
| ALD | 0 | 0.3 | 0.6 | 0.4 | 3.4 |
| SLD: PDFF ≥ 5.56% | 8.6 | 5.5 | 24.3 | 54.9 | 70.8 |
| MASLD | 6.8 | 4.6 | 22.1 | 52.1 | 66.7 |
| MetALD | 1.8 | 0.6 | 1.6 | 2.4 | 0.7 |
| ALD | 0 | 0.2 | 0.6 | 0.4 | 3.4 |
| SLD: PDFF ≥ 6.4% | 8.6 | 4.5 | 19.8 | 48.2 | 64.7 |
| MASLD | 6.8 | 3.7 | 17.9 | 45.8 | 60.6 |
| MetALD | 1.8 | 0.5 | 1.4 | 2.1 | 0.7 |
| ALD | 0 | 0.2 | 0.5 | 0.3 | 3.4 |

**Supplementary Table S6:** Men: Prevalence of steatotic liver disease and its subcategories by BMI category

| | underweight | normal weight | overweight | Obesity class 1/2 | Obesity class 3 |
|---|---|---|---|---|---|
| % of total sample | 0.9 | 35.7 | 41.8 | 20 | 1.6 |
| SLD: PDFF ≥ 5.75% | 18.7 | 13.2 | 44.2 | 75.3 | 85.6 |
| MASLD | 18.4 | 10.8 | 37.1 | 65 | 82.5 |
| MetALD | 0.2 | 1.2 | 4.7 | 7.2 | 2.1 |
| ALD | 0 | 1.3 | 2.3 | 3.2 | 1.1 |
| SLD: PDFF ≥ 5% | 18.7 | 10.8 | 40.2 | 72.1 | 83.9 |
| MASLD | 18.4 | 8.9 | 33.7 | 62.1 | 80.8 |
| MetALD | 0.2 | 1 | 4.2 | 6.9 | 2.1 |
| ALD | 0 | 0.9 | 2.2 | 3.1 | 1.1 |
| SLD: PDFF ≥ 5.56% | 18.7 | 10.2 | 38.9 | 71.3 | 82.4 |
| MASLD | 18.4 | 8.5 | 32.6 | 61.5 | 79.3 |
| MetALD | 0.2 | 0.9 | 4.1 | 6.7 | 2.1 |
| ALD | 0 | 0.8 | 2.1 | 3.1 | 1.1 |
| SLD: PDFF ≥ 6.4% | 18.4 | 8.5 | 35 | 66.8 | 76.8 |
| MASLD | 18.4 | 6.9 | 29.2 | 57.6 | 73.7 |
| MetALD | 0 | 0.8 | 3.8 | 6.3 | 2.1 |
| ALD | 0 | 0.8 | 1.9 | 3 | 1.1 |

### Supplementary Figures

**Supplementary Figure S1:** Effects of weighting on the distribution of age, sex, and socioeconomic status in the sample.

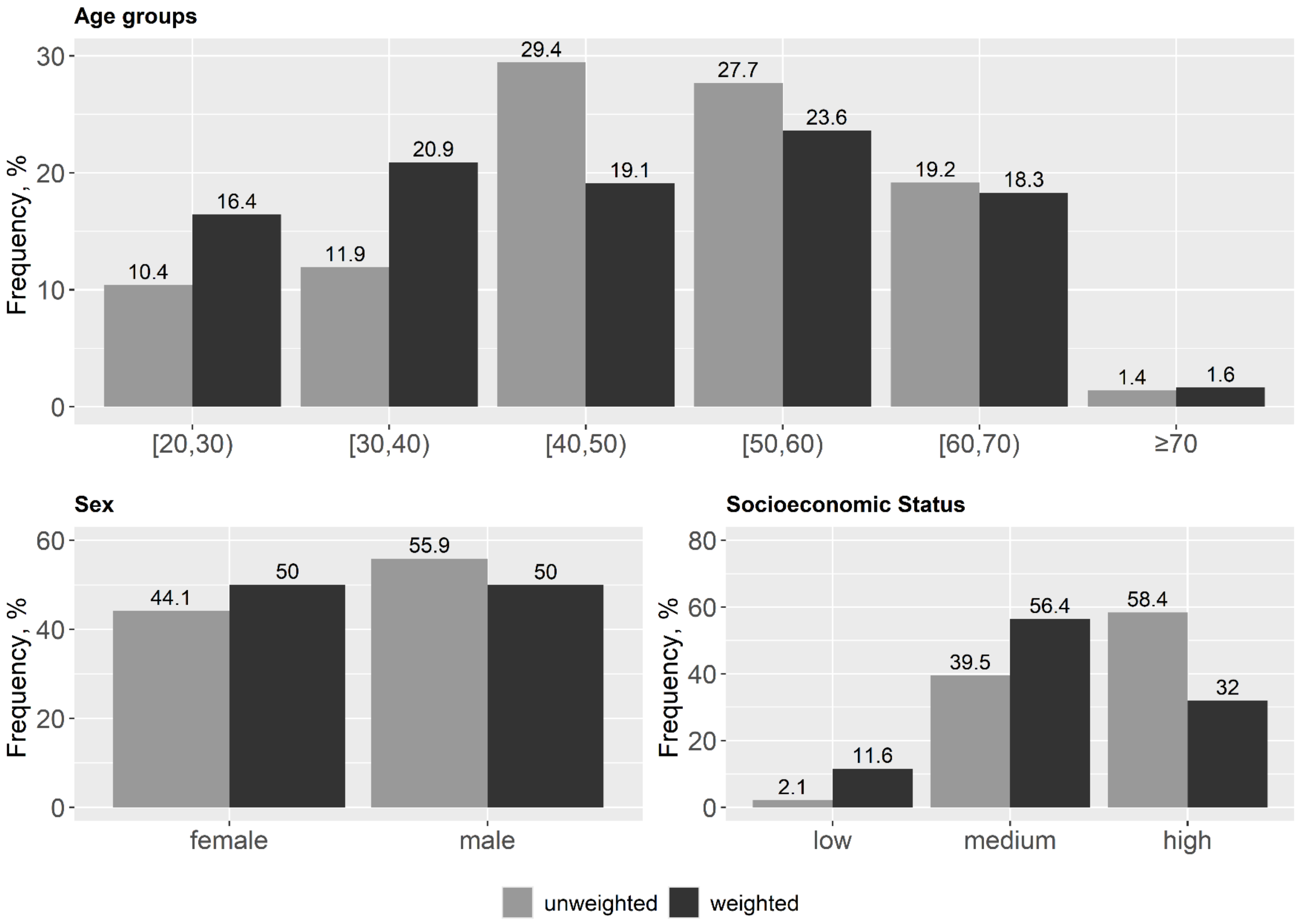


Figure shows unweighted sample frequencies (grey) and weighted population prevalences (black).

**Supplementary Figure S2**: Prevalence of SLD (steatotic liver disease), defined as PDFF ≥ 5%.

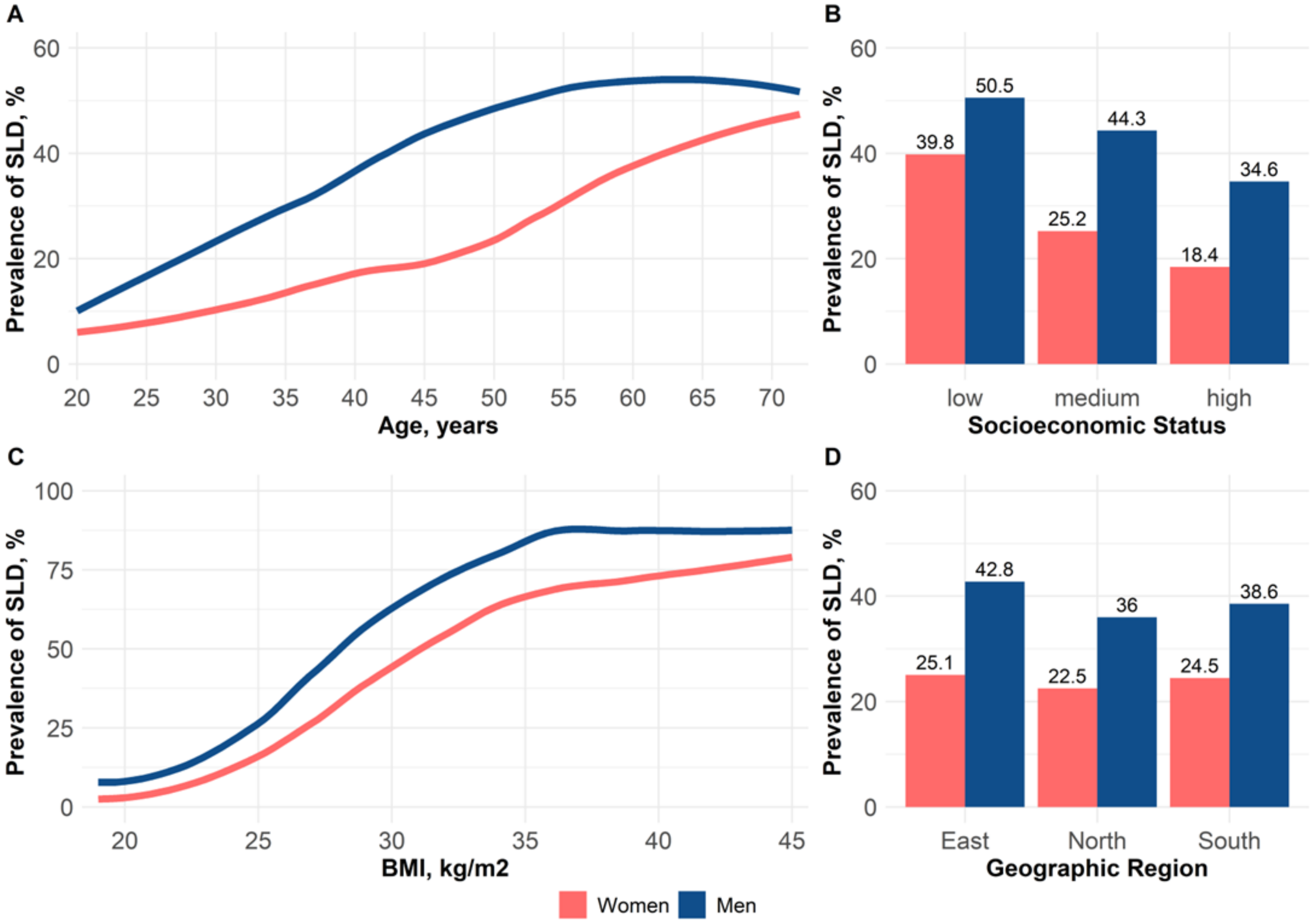

**Supplementary Figure S3:** Prevalence of SLD (steatotic liver disease), defined as PDFF ≥ 5.56%.

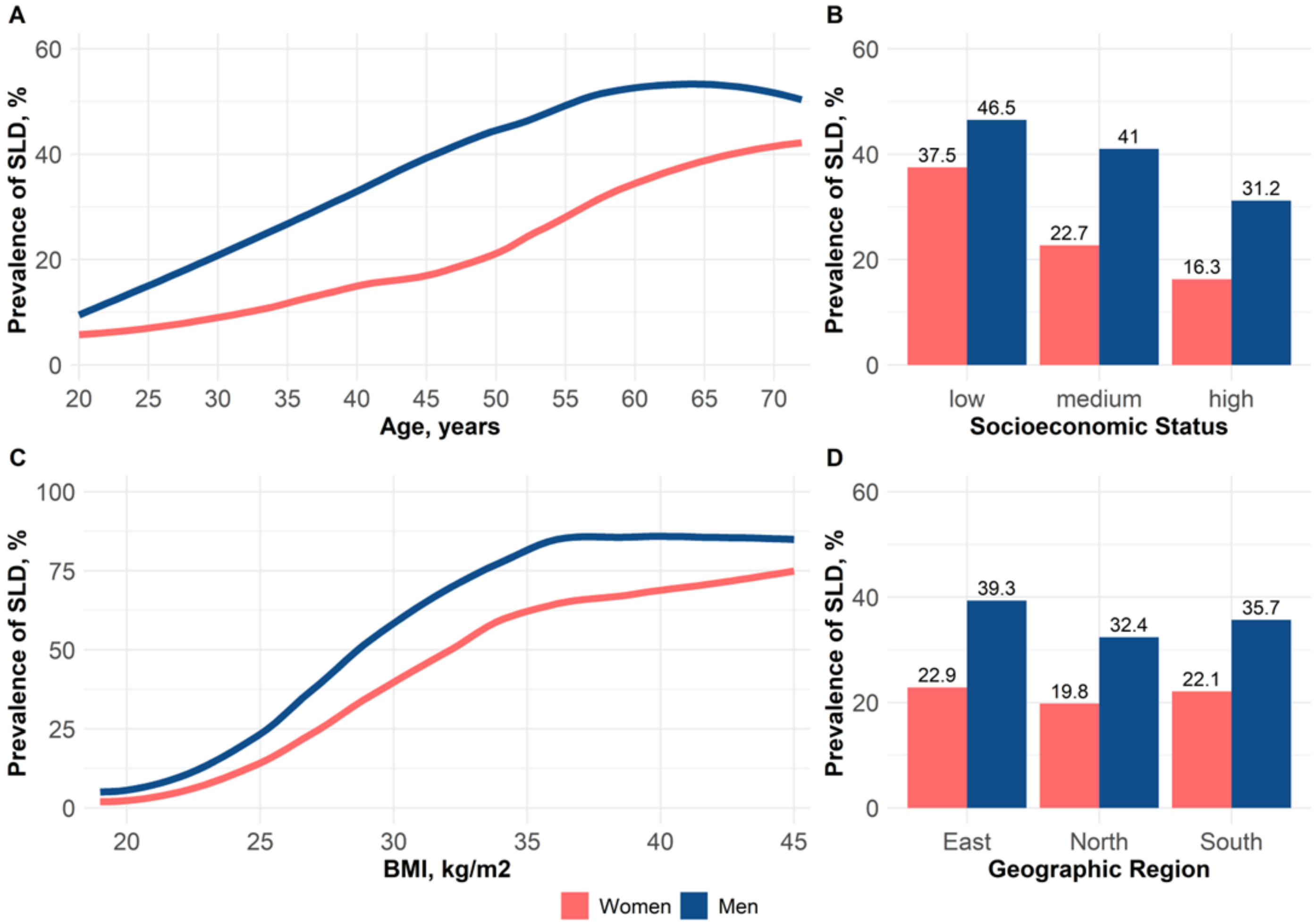

**Supplementary Figure S4:** Prevalence of SLD (steatotic liver disease), defined as PDFF ≥ 6.4%.

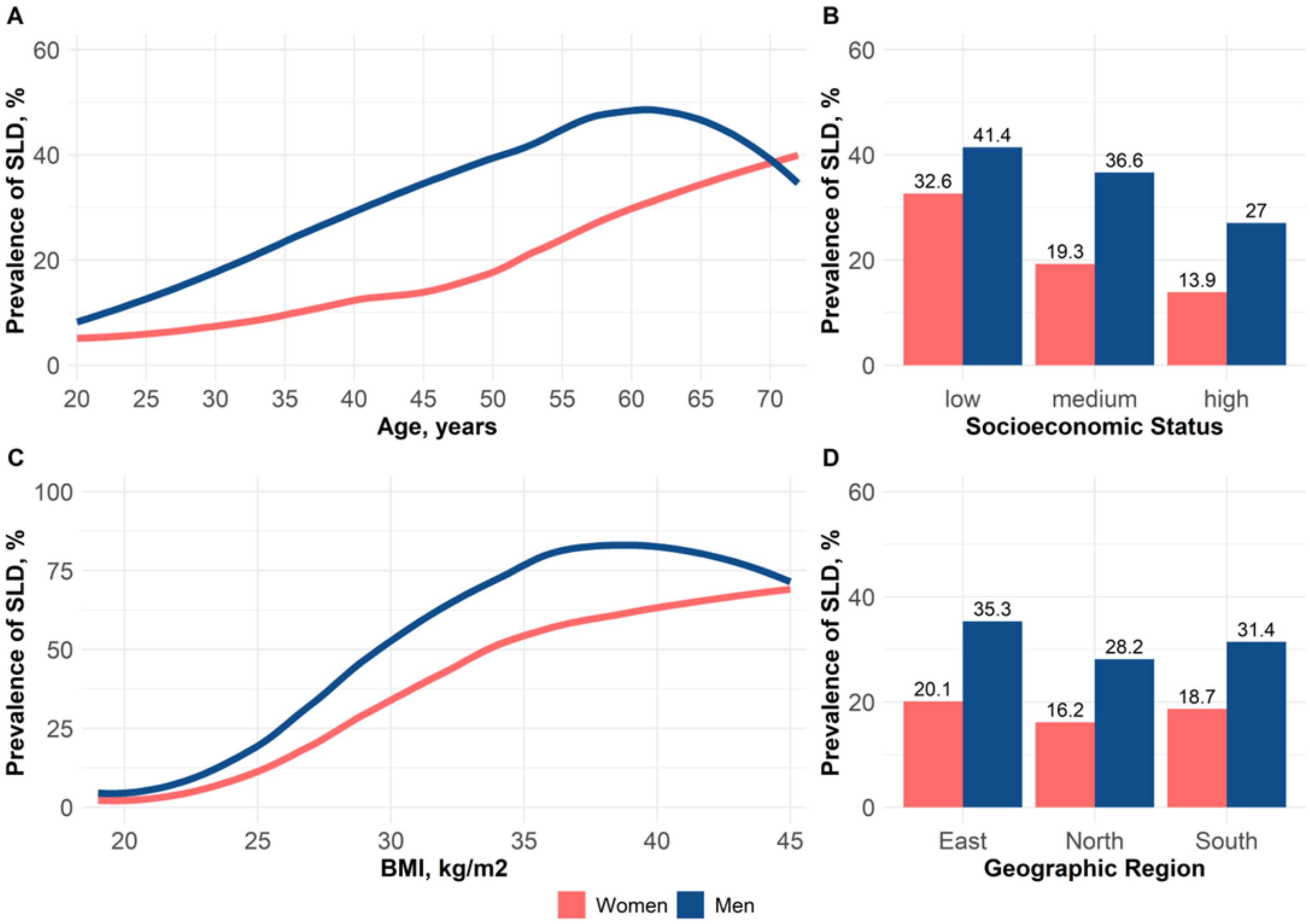

**Supplementary Figure S5:** Hepatic iron according to SLD subcategories.

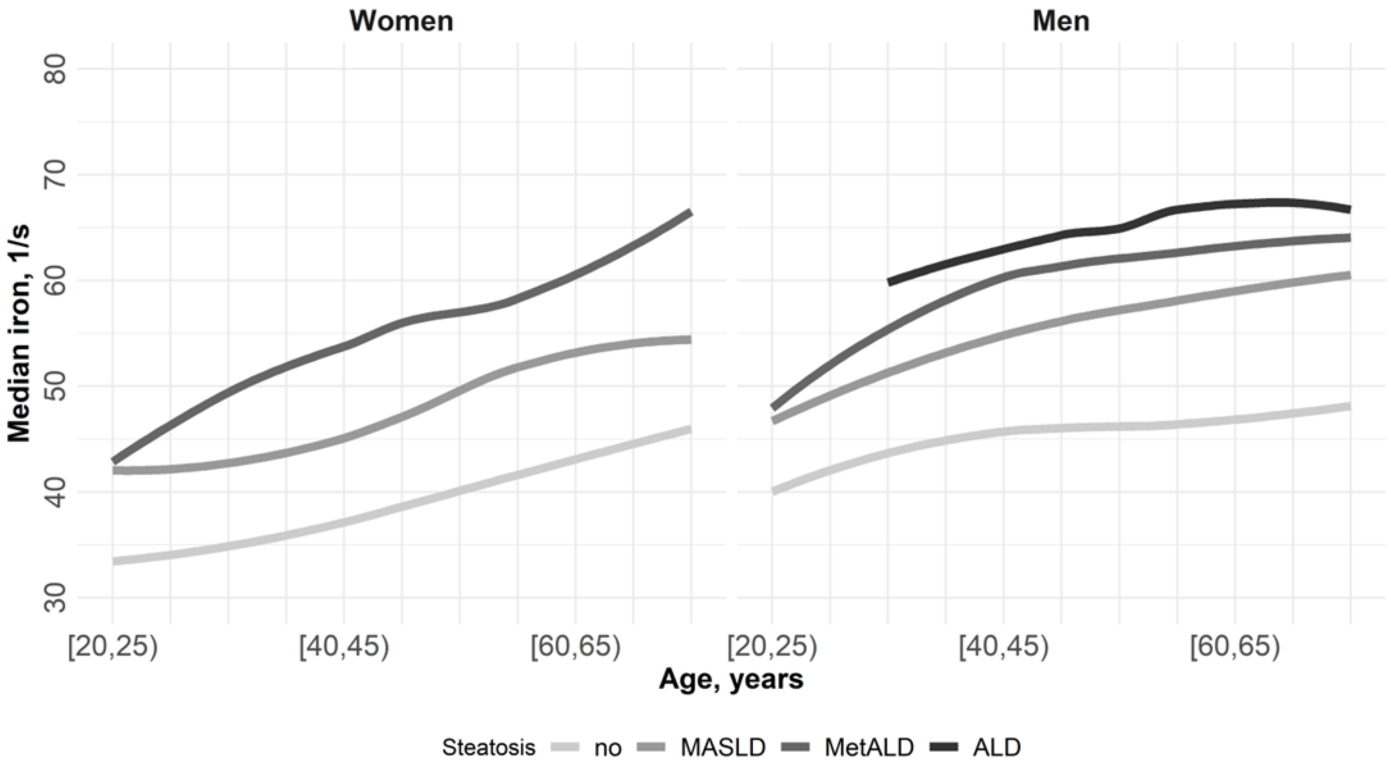


Subcategory ALD for women not shown due to low prevalence.
